# Real-time dynamic polygenic prediction for streaming data

**DOI:** 10.1101/2024.07.12.24310357

**Authors:** Justin D. Tubbs, Yu Chen, Rui Duan, Hailiang Huang, Tian Ge

## Abstract

Polygenic risk scores (PRSs) are promising tools for advancing precision medicine. However, existing PRS construction methods rely on static summary statistics derived from genome-wide association studies (GWASs), which are often updated at lengthy intervals. As genetic data and health outcomes are continuously being generated at an ever-increasing pace, the current PRS training and deployment paradigm is suboptimal in maximizing the prediction accuracy of PRSs for incoming patients in healthcare settings. Here, we introduce real-time PRS-CS (rtPRS-CS), which enables online, dynamic refinement and calibration of PRS as each new sample is collected, without the need to perform intermediate GWASs. Through extensive simulation studies, we evaluate the performance of rtPRS-CS across various genetic architectures and training sample sizes. Leveraging quantitative traits from the Mass General Brigham Biobank and UK Biobank, we show that rtPRS-CS can integrate massive streaming data to enhance PRS prediction over time. We further apply rtPRS-CS to 22 schizophrenia cohorts in 7 Asian regions, demonstrating the clinical utility of rtPRS-CS in dynamically predicting and stratifying disease risk across diverse genetic ancestries.

## Introduction

Polygenic risk scores (PRSs) – a summary of the genetic predisposition to a human complex trait or common disease across the genome – hold the promise to advance precision medicine by improving diagnostic accuracy, preventive strategies, risk stratification, and the prediction of therapeutic outcomes^1,2^. Recent methodological advancements have led to increased accuracy in polygenic prediction, both within and across populations^3–13^, facilitating the translation of PRSs from research to clinical settings^14–18^.

While promising for clinical implementation, current PRS construction methods rely on summary statistics derived from genome-wide association studies (GWASs). However, GWASs are typically conducted at intervals, often after several years and the accumulation of tens of thousands of additional samples. As a result, newly collected samples after the data freeze of a GWAS do not contribute to polygenic prediction until the next wave of GWAS is conducted. This delay implies that individuals requiring genetic risk estimation may not benefit from potential improvements in prediction accuracy that could be achieved by incorporating data from the intervening samples. The current PRS training and deployment paradigm is thus suboptimal for delivering the most accurate genetic risk prediction to incoming patients in healthcare settings, where genetic data and health outcomes are continuously being collected at an ever-increasing pace. As efforts to integrate PRS into routine clinical care continue to expand^19,20^, methods for integrating massive streaming data into PRS construction are needed to dynamically enhance prediction models over time and maximize prediction accuracy for incoming patients.

Here, building on the PRS-CS framework^6^, we introduce real-time PRS-CS (rtPRS-CS), which enables online, dynamic refinement and calibration of PRS as each new sample is collected, without the need to perform intermediate GWASs. This allows for the calculation of a PRS that leverages all available data at the time a risk prediction is needed, while avoiding the computationally intensive process of performing an updated GWAS and re-training variant weights for PRS construction. We conduct extensive simulation studies to evaluate the performance of rtPRS-CS across various genetic architectures and training sample sizes. We further apply rtPRS-CS to an array of physical measures and biomarkers that are available in both the Mass General Brigham Biobank (MGBB)^21^ and the UK Biobank (UKBB)^22,23^. Lastly, we demonstrate the clinical utility of rtPRS-CS in dynamically predicting and stratifying schizophrenia risk across diverse cohorts collected by the Stanley Global Asia Initiatives in Asian regions.

## Results

### Overview of rtPRS-CS and the study design

rtPRS-CS is an extension of PRS-CS^6^, a Bayesian method that assigns a continuous shrinkage prior on SNP effect sizes for PRS construction. Given summary statistics derived from a baseline training GWAS, PRS-CS-auto, a fully-Bayesian implementation of PRS-CS, is first applied to generate an initial estimate of posterior SNP effects, along with the global and local shrinkage parameters. For each newly arrived sample with both phenotypic and genetic information, rtPRS-CS then uses stochastic gradient descent (SGD)^24^ to iteratively refine the SNP weights, adjusting for the influences of a set of covariates, such as age, sex and genetic principal components (PCs), on the phenotype, which are estimated in an independent validation dataset (Methods). The algorithm is highly computationally efficient, taking less than 0.5 second on a single processing thread to analyze data from a new sample. As a result, the updated SNP weights can be promptly delivered to the next incoming individual in need of polygenic risk estimation. This stands in contrast to the current PRS deployment paradigm, where the same set of SNP weights are applied to all subsequent samples until the next wave of GWAS is completed and SNP weights are recalculated. Figure 1 illustrates the workflow of rtPRS-CS in real-world settings. Throughout this paper, we use a simplified scheme to evaluate rtPRS-CS, assuming that the phenotype of interest is measured for each incoming individual, which allows that sample to be incorporated into the training dataset immediately after a PRS is calculated (Supplementary Figure 1). This approach maximizes the sample size for evaluation and ensures that prediction accuracy remains unbiased, as each time a PRS is delivered, the target sample is not part of the training dataset.

**Figure 1:**
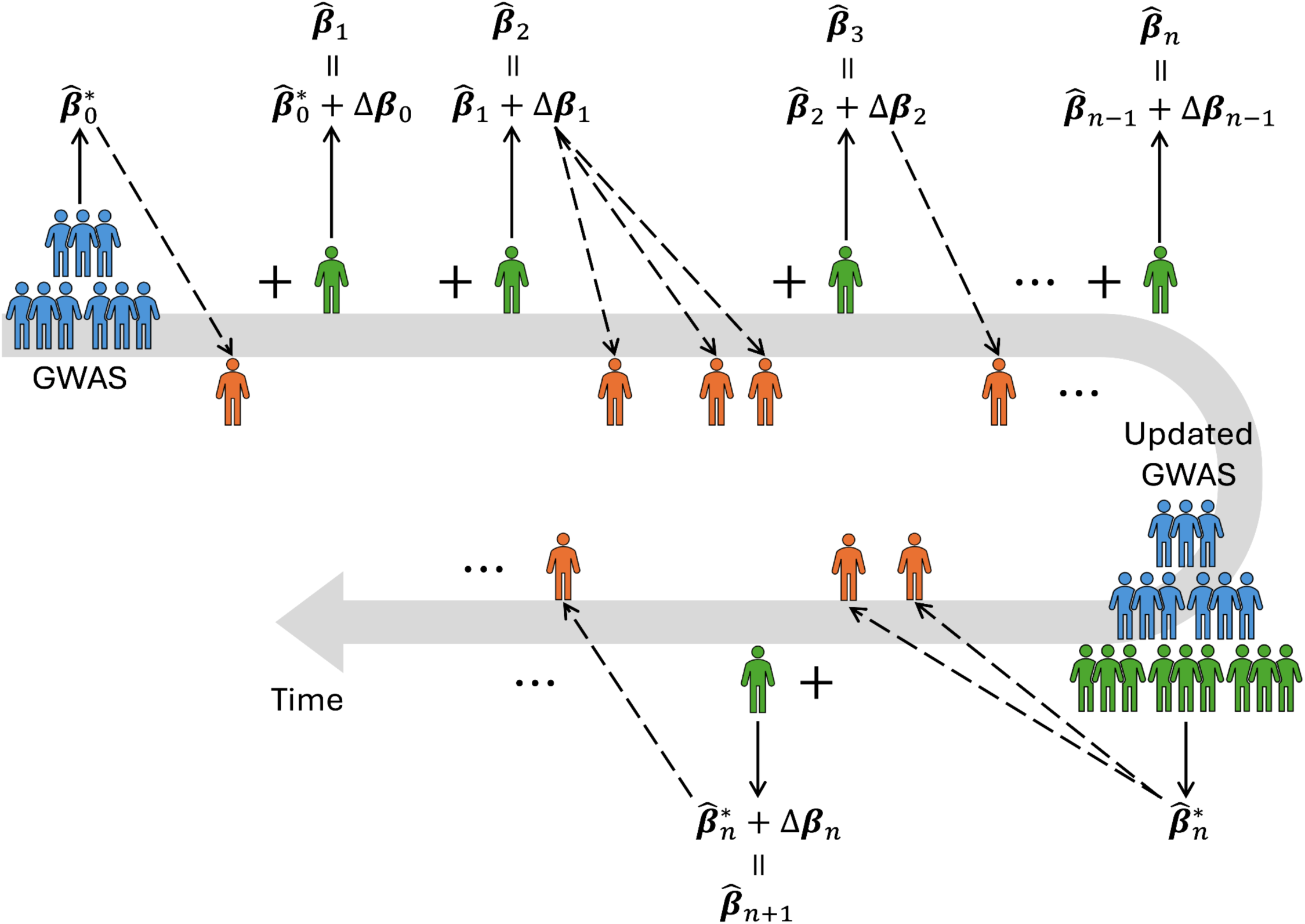
Overview of rtPRS-CS in real-world settings. Starting with the summary statistics derived from a baseline training GWAS, PRS-CS-auto is first applied to generate an initial estimate of SNP weights, denoted as 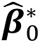, along with the global and local shrinkage parameters. Subsequently, for each newly arrived sample contributing genetic data and the phenotype of interest (shown in green), rtPRS-CS is used to update the SNP weights. For each newly arrived sample requiring polygenic risk estimation without the phenotype of interest measured (shown in orange), the latest SNP weights are utilized to calculate the PRS. Upon conducting an updated GWAS, PRS-CS-auto is applied to generate a new set of SNP weights, denoted as 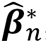, along with hyper-parameters, which serve as new starting values for running rtPRS-CS on incoming samples. A simplified scheme that was used to evaluate rtPRS-CS in this work is shown in Supplementary Figure 1.

As new samples are integrated into the training dataset to refine SNP weights, the PRS distribution may drift over time, which depends on the similarity of sample characteristics between the target and the baseline GWAS datasets. Furthermore, systematic shifts in the PRS distribution may arise among (sub)populations within the target sample due to differences in allele frequencies and linkage disequilibrium (LD) patterns. Therefore, building on existing ancestry adjustment techniques^19,25,26^, we additionally developed a regression-based method that enables dynamic calibration of individualized polygenic risk estimates across genetic ancestries and accounts for potential temporal variations in sample characteristics (Methods).

### Simulations

We first assessed the performance of rtPRS-CS via simulations. We simulated individual-level genotypes for HapMap3 variants^27^ with minor allele frequency (MAF) >1% using HAPGEN2^28^ and the 1000 Genomes Project (1KGP)^29^ phase 3 European samples (*N* = 503) as the reference panel. In our primary simulation setting, we randomly selected 1% of HapMap3 variants to be causal (i.e., polygenicity 𝜋 = 1%), which in aggregation explained 50% of phenotypic variation (i.e., SNP heritability *h*^2^ = 50%). We included 50,000 individuals in the baseline training GWAS for the initial estimation of the SNP weights. Subsequently, a target sample of 50,000 individuals independent of the GWAS cohort was assumed to become sequentially available. For each incoming target individual, a PRS was calculated using SNP weights derived from prior samples. The phenotypic and genetic data of the target individual was then used to update the SNP weights using rtPRS-CS, which were deployed to the next target individual (Supplementary Figure 1). This simulation process was repeated 20 times.

We assessed the accuracy of the dynamically refined PRS across 10 sequential bins of the target sample, each containing 5,000 individuals. As shown in Figure 2 (left panel), the prediction accuracy, measured by the squared correlation between the observed phenotype and the PRS (*R*^2^) within each bin, steadily increased as more samples were incorporated to refine the SNP weights (Supplementary Table 1). In contrast, utilizing PRS constructed from the baseline GWAS for subsequent samples yielded stable prediction accuracy across the 10 bins without notable improvement, as anticipated. To further benchmark the performance of rtPRS-CS, we compared three PRSs in a holdout sample of 50,000 individuals that was independent of the baseline and target samples: (i) PRS constructed from the baseline GWAS using PRS-CS-auto, representing the current practice for PRS construction and the performance lower bound of rtPRS-CS; (ii) PRS estimated by rtPRS-CS at the end of the training process (i.e., after refining the weights with all target samples); and (iii) PRS derived from a GWAS conducted on the combined baseline and target samples using PRS-CS-auto, representing the theoretical upper bound of the performance of rtPRS-CS. As shown in Figure 2 (right panel), the accuracy of rtPRS-CS in the holdout sample was substantially better than the lower bound and was, on average, only 1.9% lower in *R*^2^ compared to the upper bound (Supplementary Table 2). One potential factor contributing to this slightly diminished performance of rtPRS-CS relative to the theoretical upper bound could be the suboptimal estimates of the global and local shrinkage parameters derived from the baseline GWAS. These parameters were considered fixed in rtPRS-CS and were not dynamically updated alongside SNP weights, which may affect the prediction accuracy. We explored methods to overcome this constraint in the biobank analysis (see below).

**Figure 2:**
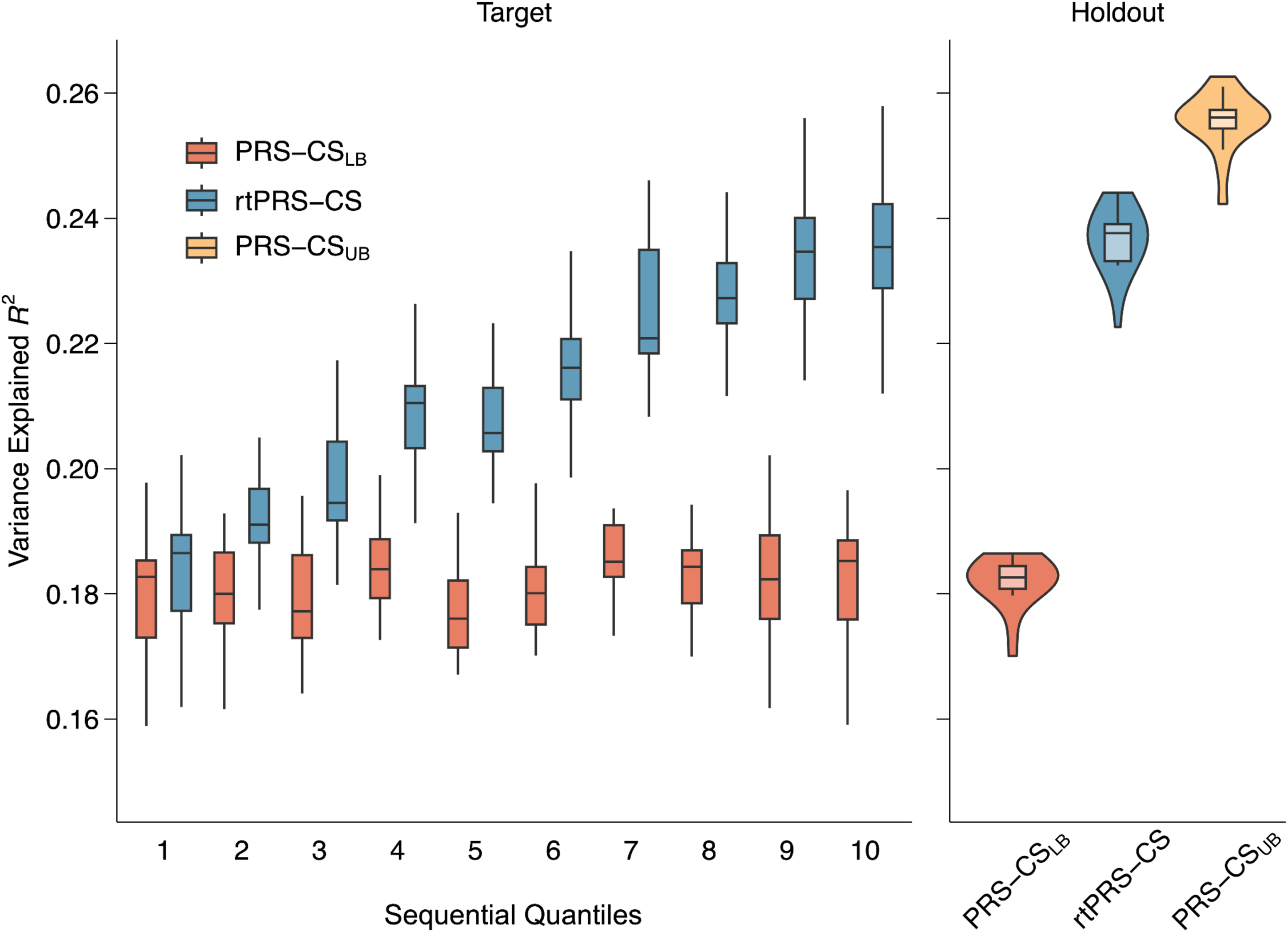
The performance of rtPRS-CS in the primary simulation setting. Left: The prediction accuracy, measured by variance explained, of the baseline PRS (red) and rtPRS-CS (blue) across 10 sequential bins of the target sample. Right: The prediction accuracy of three PRSs in the holdout sample: (i) PRS constructed from the baseline GWAS (red), representing the performance lower bound of rtPRS-CS; (ii) PRS estimated by rtPRS-CS at the end of the training process, after refining SNP weights with all target samples (blue); and (iii) PRS derived from the meta-GWAS combining baseline and target samples (yellow), representing the theoretical upper bound of the performance of rtPRS-CS. In all box plots, the middle line indicates the median across the 20 simulation replicates, and the upper and lower bound of the box indicates the 75th and 25th percentiles, respectively.

We performed additional simulations to assess the robustness of rtPRS-CS under different baseline GWAS sample sizes (*N* = 25,000 and 100,000), SNP heritability (*h*^2^ = 20% and 80%) and polygenicity (𝜋 = 0.1% and 10%). As expected, the prediction accuracy increased more rapidly with smaller baseline GWAS sample sizes. Conversely, with a baseline GWAS of a decent sample size, more subsequent samples need to be integrated before the initial PRS can be significantly improved (Supplementary Figures 2 & 3; Supplementary Tables 1 & 2). In addition, we observed that the predictive performance of rtPRS-CS approached its theoretical upper bound for phenotypes with a more polygenic architecture (i.e., lower heritability and higher polygenicity).

Conversely, for phenotypes with a sparser genetic architecture (i.e., higher heritability and lower polygenicity), the upper bound was notably higher than the accuracy that rtPRS-CS was able to achieve (Supplementary Figures 4-7; Supplementary Tables 1 & 2). This suggests that phenotypes that are easier to predict may benefit more from accurate estimation of shrinkage parameters in the rtPRS-CS model. Nevertheless, in all simulation settings, rtPRS-CS demonstrated continuous improvement in prediction accuracy as the number of contributing samples increased, suggesting that rtPRS-CS is robust to varying genetic architectures and baseline training sample sizes.

### Quantitative trait prediction in biobanks

Next, we assessed the performance of rtPRS-CS across 21 quantitative traits collected by both the Mass General Brigham Biobank (MGBB)^21^ and the UK Biobank (UKBB)^22,23^. MGBB served as the baseline training dataset. We conducted a GWAS of each trait within the MGBB sample, with sample sizes ranging from 10,475 to 28,817 (Supplementary Table 3), and applied PRS-CS-auto^6^ to obtain initial estimates of SNP weights, along with estimates of global and local shrinkage parameters. The target dataset comprised up to 300,054 UKBB samples of European ancestries (Supplementary Table 4), assumed to arrive sequentially in a randomized order. We iteratively applied the latest SNP weights, derived from all prior samples, to an incoming individual to generate a polygenic prediction, and subsequently used the phenotypic and genetic data from that individual to update the SNP weights using rtPRS-CS (Supplementary Figure 1). Similar to the simulation studies, we assessed the predictive performance of rtPRS-CS using variance explained (*R*^2^) across 10 sequential bins of the UKBB target sample, adjusting for age, sex and top genetic PCs. Figure 3 and Supplementary Figures 8-10 show that, for all traits, the dynamically updated PRS estimated using rtPRS-CS quickly surpassed the performance of the PRS constructed from the baseline GWAS, after 10% of the target samples were incorporated, and continued to improve across sample quantiles (Supplementary Table 5).

**Figure 3:**
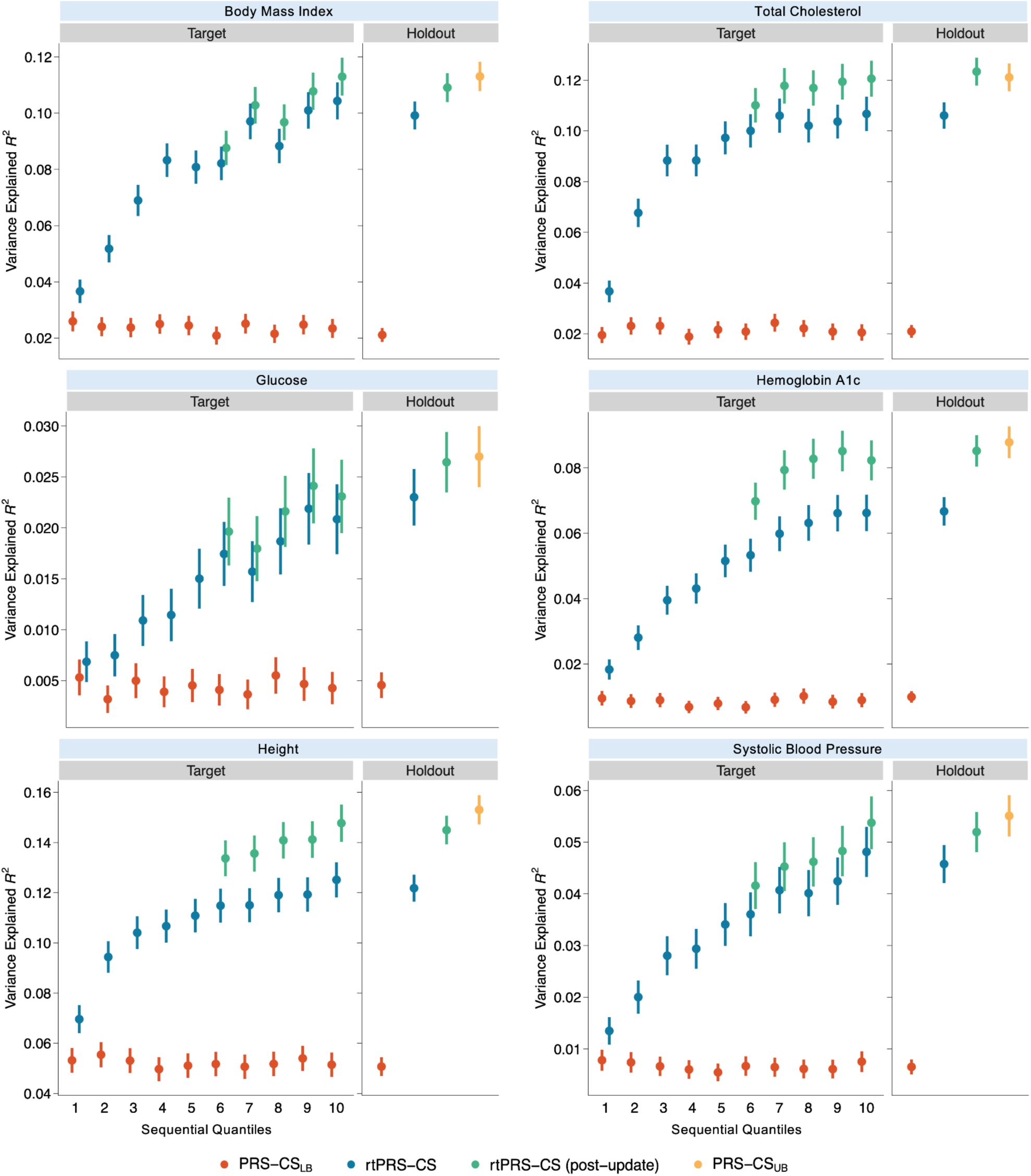
The performance of rtPRS-CS for representative quantitative traits in biobanks. For each trait, Left: The prediction accuracy, measured as variance explained, of the baseline PRS (red), rtPRS-CS (blue), and rtPRS-CS with halfway training GWAS update (green), across 10 sequential bins of the UK Biobank target sample. Right: The prediction accuracy of four PRSs in the UK Biobank holdout sample: (i) PRS constructed from the baseline GWAS (red), representing the performance lower bound of rtPRS-CS; (ii) PRS estimated by rtPRS-CS at the end of the training process, after refining SNP weights with all target samples (blue); (iii) PRS estimated by rtPRS-CS with halfway update of the GWAS (green); and (iv) PRS derived from the meta-GWAS combining baseline and target samples (yellow), representing the theoretical upper bound of the performance of rtPRS-CS. In all plots, the error bars represent 95% confidence intervals.

We compared the predictive performance of three PRSs in a holdout sample of 50,000 UKBB individuals, which was independent of the training and target datasets: (i) PRS derived from applying PRS-CS-auto to the baseline MGBB GWAS, representing the current practice for PRS construction and the performance lower bound of rtPRS-CS; (ii) PRS estimated by rtPRS-CS at the end of the training process (i.e., after refining the weights with all target UKBB samples); and (iii) PRS derived from applying PRS-CS-auto to the fixed-effect meta-analysis of the MGBB baseline GWAS and the UKBB target sample GWAS, representing the theoretical upper bound of rtPRS-CS performance. While rtPRS-CS significantly outperformed the baseline PRS, its predictive performance for many traits was close, but notably lower than the theoretical upper bound, especially for traits whose baseline GWAS was underpowered (Supplementary Table 6). This performance gap can likely be attributed to inaccurate shrinkage parameter estimates derived from the small MGBB baseline GWAS. We therefore explored the benefits of periodically updating the GWAS and the estimates of shrinkage parameters. Specifically, after half (five quantiles) of the UKBB target samples were incorporated, we conducted an intermediate meta-analysis of the MGBB baseline GWAS and the UKBB GWAS using the first half of the samples. Subsequently, PRS-CS-auto^6^ was applied to the meta-GWAS to derive SNP weights and update the estimates of global and local shrinkage parameters. These updated parameters were then used as a new set of starting values for running rtPRS-CS on the remaining UKBB target samples. Figure 3 and Supplementary Figures 8-10 show that, with the intermediate update, rtPRS-CS yielded prediction accuracy that was statistically indistinguishable from the theoretical upper bound for most traits in the holdout sample (Supplementary Table 6). Overall, our biobank analysis demonstrated that rtPRS-CS can dynamically enhance the accuracy of PRS across traits with varying genetic architectures when transferring a baseline PRS to a target sample with distinct sample characteristics (e.g., from healthcare system-based biobanks such as MGBB to population-based biobanks such as UKBB).

### Schizophrenia risk prediction

Lastly, we evaluated the performance of rtPRS-CS in schizophrenia risk prediction across 22 cohorts of Asian ancestries (Supplementary Table 7), with a total of 27,159 schizophrenia cases and 32,336 controls, collected by the Stanley Global Asia Initiatives. The baseline GWAS consisted of a meta-analysis of five cohorts, comprising 4,343 cases and 7,957 controls, for which only GWAS summary statistics were available. Two cohorts, JPN1 and KOR1, were set aside as holdout testing datasets. Individual-level data from the remaining 15 cohorts were merged, and the order of the merged samples was randomized. We randomly selected 1,000 cases and 1,000 controls to form the validation dataset, where the effects of sex and genetic PCs on schizophrenia risk were estimated. The remaining 21,581 cases and 23,347 controls constituted the target sample and were assumed to arrive sequentially. For each incoming sample, the process of PRS calculation using weights trained on prior samples followed by updating SNP weights using rtPRS-CS was iteratively performed.

The schizophrenia dataset included samples from various Asian regions, including Mainland China, Hong Kong, Taiwan, Japan, Korea, Singapore and Indonesia, with diverse genetic ancestries (Figure 4a; Supplementary Table 7). Raw PRS estimated by PRS-CS-auto or rtPRS-CS exhibited major distributional shifts by cohort as anticipated, particularly noticeable for Japanese and Indonesian samples (Figure 4b), which are genetically distant from other samples (Figure 4a). Hence, without accounting for population structure within the dataset, estimated risks were not comparable across cohorts, potentially leading to decreased prediction accuracy and patient misclassification. Following the dynamic calibration of the PRS for each incoming sample against the PRS distribution of preceding control samples (Methods), individualized polygenic risk estimates can be standardized on the same scale across cohorts in real time as SNP weights are updated (Figure 4b).

**Figure 4:**
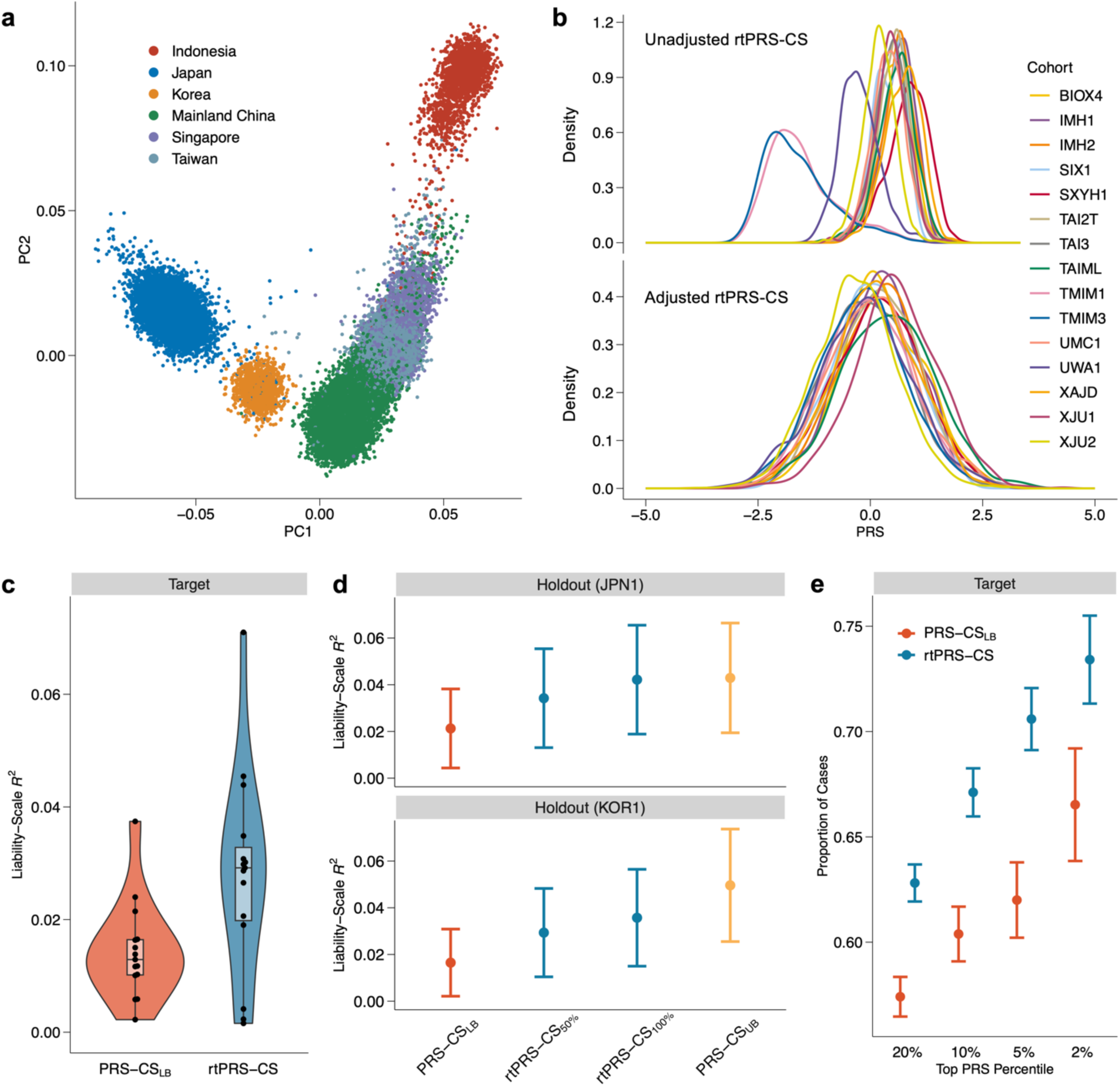
The performance of rtPRS-CS in the schizophrenia analysis. **a**, The top two genetic principal components (PCs) of schizophrenia cases and controls with individual-level data. **b**, The distribution of PRSs estimated by rtPRS-CS by cohort, with (lower panel) and without (upper panel) dynamic regression-based adjustment to account for population structure. **c**, The prediction accuracy, measured by variance explained on the liability scale, of the baseline PRS (red) and rtPRS-CS (blue) in each target schizophrenia cohort. In the box plot, the middle line indicates the median prediction accuracy across cohorts, and the upper and lower bound of the box indicates the 75th and 25th percentiles, respectively. **d**, The prediction accuracy, measured by variance explained on the liability scale, of four PRSs in the two holdout cohorts (JPN1 and KOR1): (i) PRS derived from the baseline GWAS, representing the performance lower bound of rtPRS-CS; (ii) PRS estimated by rtPRS-CS at halfway through the training process; (iii) PRS estimated by rtPRS-CS at the end of the training process; and (iv) PRS derived from the meta-GWAS of the baseline and full target sample, representing the theoretical upper bound of rtPRS-CS performance. **e**, The proportion of schizophrenia cases at different risk cutoffs, identified by the baseline PRS (red) and rtPRS-CS (blue), across all target cohorts. In **d** and **e**, the error bars represent 95% confidence intervals.

After generating and calibrating PRS for each target sample using rtPRS-CS, we assigned samples back to their respective contributing cohorts, and examined the prediction accuracy within each cohort, adjusting for sex and in-sample PCs. This ensured that the performance evaluation was minimally influenced by population stratification. Across the 15 cohorts that contributed to the target dataset, rtPRS-CS improved the median variance explained in schizophrenia liability from 1.3%, as observed when applying the PRS trained on the baseline GWAS, to 2.9% (Figure 4c; Supplementary Table 8). In the two holdout cohorts, rtPRS-CS also demonstrated notable improvement in out-of-sample prediction accuracy compared to the baseline PRS (Figure 4d; Supplementary Table 9). Moreover, the PRS trained by rtPRS-CS after integrating all target samples exhibited prediction accuracy comparable to the PRS trained directly on the meta-GWAS across baseline and target cohorts, representing the theoretical performance upper bound, although evaluation metrics had large estimation uncertainties due to sample size limitations.

Finally, we examined the capability of rtPRS-CS in identifying individuals at high risk of schizophrenia. Unified thresholds, corresponding to the top 20%, 10%, 5% and 2% of schizophrenia risk among the control group, were applied to the calibrated PRS for each target sample (Methods). Figure 4e shows that, across target cohorts, rtPRS-CS identified significantly more schizophrenia cases compared to PRS constructed from the baseline GWAS (Supplementary Table 10). For instance, among individuals in the top 2% of the rtPRS-CS distribution, 73.4% were schizophrenia patients, making a 1.68-fold increase in the proportion of cases relative to middle quantiles (40%-60%) of the distribution, compared to 66.5% cases in the top 2% of the baseline PRS. Overall, the schizophrenia analysis showed that rtPRS-CS, including dynamic updating of SNP weights and dynamic PRS calibration, can improve disease risk prediction and stratification in clinical settings with samples from diverse genetic backgrounds.

## Discussion

We introduced rtPRS-CS, a method designed to incorporate streaming data for updating SNP weights in real-time, rather than at sporadic intervals, thereby maximizing the prediction accuracy of PRS for incoming samples. Through extensive simulations, we showed that rtPRS-CS dynamically improves predictive performance across traits with varying genetic architectures. Using quantitative physical measures and biomarkers from biobanks, we showed that rtPRS-CS enhances the accuracy of PRS over time, even when the initial PRS was trained on datasets with different sample characteristics compared to the target individuals. The analysis of schizophrenia cohorts from various Asian regions further demonstrated that rtPRS-CS can dynamically calibrate disease risk prediction when applied to samples with heterogeneous genetic ancestries and collected using different genotyping platforms.

rtPRS-CS relies on stochastic gradient descent (SGD)^24^ – a cornerstone algorithm used for training and optimizing a wide range of deep learning models^30^ – to dynamically update SNP weights as each new sample is collected. A number of potential enhancements to the rtPRS-CS algorithm can thus be explored by incorporating advancements from research in the machine learning field. For example, rtPRS-CS does not update global and local shrinkage parameters over time, which may limit prediction accuracy, especially for traits with underpowered baseline GWASs. That said, this approach facilitates a stable and highly computationally efficient closed-form update of SNP weights, and we have shown that the limitation of fixing shrinkage parameters can be largely mitigated by periodically performing intermediate GWASs and updating their estimates. Future research could explore integrating hyper-parameters into the SGD algorithm while preserving computational stability and efficiency. rtPRS-CS may also benefit from implementing the mini-batch updating scheme, which may yield less noisy updates to SNP weights compared to utilizing a single data point at a time. Additionally, incorporating the method of momentum and adaptive learning rate algorithms^31,32^ into rtPRS-CS may potentially accelerate the learning process. In particular, one interesting future direction involves up-weighting the contribution of incoming samples to expedite the adaptation of predictions to the local genetic and socio-environmental characteristics of the target population.

The clinical translation of PRSs, particularly rtPRS-CS, is confronted with several challenges. A fundamental consideration for implementing rtPRS-CS in routine healthcare is the ability to integrate data from and deliver genetic risk predictions to individuals of diverse genetic and sociocultural backgrounds. This ensures that advances in genomic medicine can benefit global populations^14,19,33,34^. In the current study, we have shown that the dynamic calibration algorithm effectively adjusts individualized polygenic risk estimates against preceding samples by removing variations in the PRS distribution that are correlated with population structure, producing well-calibrated schizophrenia risk predictions across individuals with diverse Asian ancestries. While our primary focus was on genetic ancestry adjustment, this method can be extended to model and calibrate PRS across a broader range of contexts^35^. Future research is required to assess the performance of this dynamic calibration algorithm across a wider spectrum of disease phenotypes and in datasets that capture global genetic and sociocultural diversity. While the schizophrenia cohorts captured substantial genetic variation within Asia, one limitation of rtPRS-CS is its current restriction to integrating samples from relatively homogeneous continental populations. A straightforward extension could involve combining the SGD algorithm with the PRS-CSx framework^3,25^, thereby allowing for the modeling of samples from multiple genetic ancestries.

However, a rtPRS-CSx model may remain suboptimal in clinical settings as it requires assigning individuals to discrete population groups, which can be difficult for individuals of admixed or complex genetic ancestries.

Developing polygenic prediction methods that can model global genetic variation as a continuum^36^ is challenging and represents an important future direction.

Important ethical considerations also arise when implementing rtPRS-CS in real-world clinical settings. For instance, if a binarized PRS prediction (e.g., high risk vs. average risk) were to be provided to patients, dynamic real-time updates of SNP weights could lead to conflicting predictions within a short time frame, especially for patients whose disease risk is near the clinical cutoff, thus complicating decision-making. This highlights the current imprecision of PRS for individualized disease risk prediction, particularly for minority populations that are underrepresented in genetic research, and underscores the importance of integrating uncertainty measures into PRS calculation, interpretation and communication^37^. Other practical considerations of implementing rtPRS-CS in real-world settings include how to account for related individuals. Although leveraging information on relatives of target individuals can boost prediction accuracy^38^, dynamically incorporating numerous closely related individuals into the training dataset may bias SNP weights as most PRS methods, including rtPRS-CS, were derived under linear models with the assumption of sample unrelatedness. However, in the schizophrenia analysis, which included multiple trio and multiplex family cohorts, rtPRS-CS demonstrated robust improvements over the baseline PRS. In healthcare settings, close familial relatedness often represents only a small portion of the entire patient sample and thus may have limited impact on PRS accuracy. Future research may improve the modeling of related samples by designing adaptive and optimal learning rates within the SGD algorithm. Lastly, while the genotype of an individual remains static, health conditions are highly dynamic, and diagnostic information – such as switch in diagnosis and transitioning from control to case status – can evolve over time. rtPRS-CS has the potential to better capture this time-varying information. For example, by dynamically removing or updating the contribution of an individual to the SNP weights when their diagnosis or health status changes, rtPRS-CS may further improve the accuracy of polygenic prediction compared to PRS constructed from cross-sectional GWASs. With these being said, we note that the current version of rtPRS-CS is not intended to replace conventional GWAS, which offers more rigorous harmonization and quality control of phenotypic and genetic data, as well as more accurate estimates of SNP effect sizes and model hyper-parameters. Further investigations are required to explore the optimal schedule for periodic GWAS updates as a function of the genetic architecture of the phenotype and the baseline sample size.

In summary, we have developed rtPRS-CS, a polygenic prediction method that can leverage streaming data to dynamically calibrate and refine disease risk prediction, which holds particular promise for biomarker and disease risk predictions where data from diverse populations are rapidly and continuously being generated. As PRSs become more integrated into routine healthcare, rtPRS-CS may enhance and maximize their clinical utility by delivering the most up-to-date genetic risk predictions to incoming patients.

## Methods

### rtPRS-CS

rtPRS-CS builds on the PRS-CS framework^6^. Consider a high-dimensional Bayesian linear regression model:

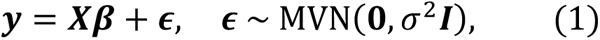

where 𝒚 is a vector of standardized phenotypes (with zero mean and unit variance) from 𝑁 individuals, 𝑿 is an 𝑁 × 𝑀 standardized genotype matrix (each column has zero mean and unit variance), 𝜷 is a vector of SNP effects, 𝝐 is a vector of normally distributed residual effects with variance 𝜎^2^, and 𝑰 is an identity matrix. We assign a continuous shrinkage prior on the effect size of each SNP, 𝛽_*j*_, which can be represented as global-local scale mixtures of normals:

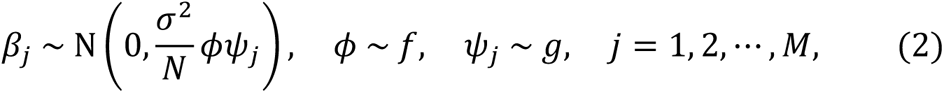

where 𝑓 and 𝑔 are density functions, 𝜙 is the global shrinkage parameter that models the overall sparseness of the genetic architecture, and 𝜓*_j_* is the local, variant-specific shrinkage parameter. Carefully designed 𝑓 and 𝑔 can shrink noise coefficients while retaining real signals. In PRS-CS-auto^6^, we assigned a gamma-gamma prior on 𝜓_*j*_ and a half-Cauchy prior on 𝜙. In general, continuous shrinkage priors allow for block update of regression coefficients, offering rapid mixing and convergence as well as robustness to LD misspecification.

Given an initial estimate of the SNP effects, denoted as 𝜷_0_, along with estimates of global and local shrinkage parameters 𝜙 and 𝜓*_j_*, we use stochastic gradient descent (SGD)^24^ to update 𝜷 ∈ ℝ*^M^* from sequentially arriving data points (𝒙*_n_*, 𝑦*_n_*), such that conditional on 𝒙*_n_* ∈ ℝ*^M^*, 𝑦*_n_* ∈ ℝ follows the distribution 𝑓(𝑦*_n_*; 𝒙*_n_*, 𝜷) specified by Eq. (1) and (2). The explicit SGD, defined for 𝑛 = 1, 2, ⋯, can be written as:

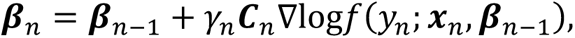

where ∇ is the gradient of a function, 𝛾*_n_* is the learning rate sequence, often defined as 𝛾*_n_* = 𝛾_1_𝑛^-𝛾^, in which 𝛾_1_ > 0 is the learning rate parameter, 𝛾 ∈ (0.5,1], and 𝑪*_n_* is a sequence of positive-definite matrices known as condition matrices. Using the same notations, the implicit SGD is defined as:

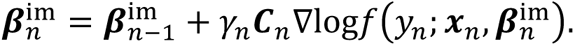

We note that in explicit SDG, the next iterate 𝜷*_n_* can be immediately computed given 𝜷*_n_*_-1_ and the new data point (𝒙*_n_*, 𝑦*_n_*), while in implicit SDG, the next iterate appears on both sides of the equation.

The log-likelihood of the model, defined by Eq. (1) and (2), for a new data point (𝒙*_n_*, 𝑦*_n_*) is (up to a constant):

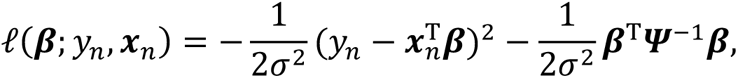

where 𝜳 = 𝜙diag{𝜓_1_, 𝜓_2_, ⋯, 𝜓*_M_*} is a diagonal matrix containing global and local shrinkage parameters. The gradient of the log-likelihood is thus given by:

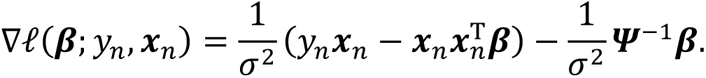

Assuming that 𝜎^2^ has been absorbed into the learning rate parameter 𝛾_*n*_, the explicit and implicit SGD procedures can be expressed as:

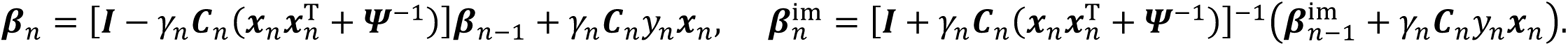

To guide the optimal selection of the learning rate parameter and the condition matrices, it can be proved, using mathematical induction, that when 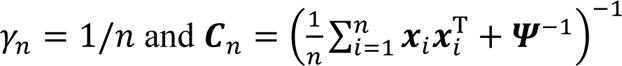, the parameter estimates obtained by the explicit SGD procedure is equivalent to the penalized least squares estimator. We further notice that 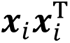, is an estimate of the LD matrix, denoted as 𝑫, and can be approximated using an external reference panel. 𝑫 can thus be treated as a constant matrix and does not need to be updated in each iterate. Therefore, the explicit and implicit SGD procedures can be further simplified as 𝑪*_n_* = (𝑫 + 𝜳^-1^)^-1^ and

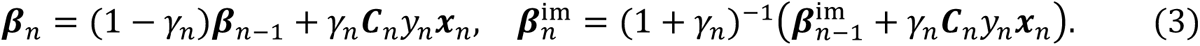

It can be seen from Eq. (3) that implicit SGD iterates are shrinked versions of explicit SGD. Consistent with our numerical experiments, recent theoretical and empirical studies have shown that implicit SDG is numerically more stable and is robust to the misspecification of the learning rate^24^. Therefore, in this work, we focused on the evaluation of the implicit SDG.

In practice, we use PRS-CS-auto^6^ to generate an initial estimate of SNP effects, along with the global and local shrinkage parameters. We use a validation dataset that is representative of the incoming target samples to estimate the scale of the phenotype, the influences of covariates on the phenotype, as well as the allele frequency of each genetic variant. For each target sample, the covariate effects are subtracted from the phenotype and the residualized phenotype and genotype are standardized before the implicit SDG algorithm is applied to update the SNP weights.

### Dynamic PRS calibration

To account for temporal shifts in sample characteristics and population structure within the target sample, we dynamically calibrate the PRS of an incoming sample against the PRS distribution of preceding samples, using a similar approach implemented in prior work^19,25,26^. Specifically, for an incoming individual 𝑖 in the target dataset, we fit the following two sequential regressions in the prior 𝑠 individuals (for disease phenotypes, we use the prior 𝑠 control samples):

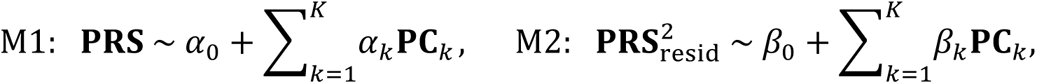

where 𝐏𝐑𝐒_resid_is the residual of M1. Fitting the two regressions M1 and M2 estimates how the mean and spread of the PRS distribution vary with population structure captured by PCs. For individual 𝑖 projected onto the same PC space with the raw score PRS*_i_*_,raw_ output by rtPRS-CS, the dynamically adjusted and calibrated PRS is calculated as:

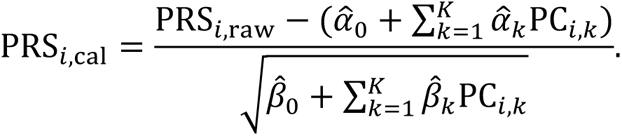

In this work, we use 𝐾=5 genetic PCs to capture broad-scale population structure.

### Simulations

Individual-level genotypes of European ancestries were simulated using HAPGEN2^28^, with 1000 Genomes Project (1KGP)^29^ phase 3 European samples (*N* = 503) as the reference panel. We constrained the analysis to HapMap3 variants^27^ with minor allele frequency (MAF) >1% in the European population. In our primary simulation setting, we randomly selected 1% of HapMap3 variants as causal (i.e., polygenicity 𝜋 = 1%), with their per-allele effect sizes drawn from a standard normal distribution. The phenotype was then generated using 𝒚 = 𝑿𝜷 + 𝝐, where 𝑿 was the genotype matrix, 𝜷 was a vector of the simulated effect sizes, and 𝝐 was a vector of normally distributed noise, such that genetic effects in aggregation explained 50% of phenotypic variation (i.e., SNP heritability *h*^2^ = 50%). This simulation process was repeated 20 times.

We included 50,000 individuals in the baseline training GWAS and 10,000 individuals in the validation dataset. PRS-CS-auto was applied to the baseline GWAS summary statistics to produce initial estimates of SNP weights along with estimates of the global and local shrinkage parameters. The validation dataset was used to estimate allele frequencies as well as the scale (standard deviation) of the phenotype, which were used to standardize the genotypes and phenotypes of the target sample. A target sample of 50,000 individuals, independent of the training and validation cohorts, was assumed to become sequentially available. For each incoming target individual, a PRS was calculated using SNP weights derived from prior samples. The phenotypic and genetic data of the target individual was then used to update the SNP weights using rtPRS-CS, which were then deployed to the next target individual (Supplementary Figure 1). rtPRS-CS was assessed across 10 sequential bins of the target sample, each containing 5,000 individuals. The prediction accuracy was measured using the squared correlation between the observed phenotype and the PRS.

We also set aside an independent holdout sample of 50,000 individuals, which was used to compare the predictive performance of three PRSs: (i) PRS constructed from the baseline GWAS using PRS-CS-auto, representing the current practice for PRS construction and the performance lower bound of rtPRS-CS; (ii) PRS estimated by rtPRS-CS at the end of the training process (i.e., after refining the weights using all target samples); and (iii) PRS derived from a meta-GWAS (combining baseline and target samples) using PRS-CS-auto, representing the theoretical upper bound of the performance of rtPRS-CS.

We conducted a series of secondary simulations to assess the robustness of rtPRS-CS under different baseline GWAS sample sizes (*N* = 25,000 and 100,000), SNP heritability (*h*^2^ = 20% and 80%) and polygenicity (𝜋 = 0.1% and 10%).

### Quantitative trait prediction in biobanks

We assessed the performance of rtPRS-CS across 21 quantitative traits measured in both the Mass General Brigham Biobank (MGBB)^21^ and the UK Biobank (UKBB)^22,23^.

MGBB is a U.S. hospital-based biobank, which collected plasma, serum, DNA and buffy coat samples from consented patients, with linkage to their electronic health records. UKBB is a prospective population-based cohort study comprising approximately 500,000 individuals recruited across Great Britain. Both datasets underwent standard genetic quality control and were restricted to individuals of European ancestries. Within MGBB, when multiple measures of a trait were available for an individual, the median value was used. Furthermore, measures falling outside 5 standard deviations from the sample mean were removed to mitigate recording errors.

A total of 31,322 MGBB samples comprised the baseline training dataset, with sample sizes ranging from 10,475 for total protein to 28,817 for calcium (Supplementary Table 3). We conducted baseline GWAS of each trait, adjusting for age, sex and top 20 genetic PCs. For lipid traits (high-density lipoprotein, low-density lipoprotein and total cholesterol), we additionally included history of statin use as a covariate. We then applied PRS-CS-auto to baseline GWAS summary statistics to obtain initial estimates of SNP weights, along with estimates of global and local shrinkage parameters.

Up to 375,054 UKBB individuals were available for analysis, with sample sizes ranging from 305,227 for direct bilirubin to 374,231 for diastolic blood pressure (Supplementary Table 4). We randomly split the UKBB dataset into a validation sample of 25,000 individuals, a holdout sample of 50,000 individuals, and a target sample of 300,054 individuals. The validation sample was used to define the UKBB PC space, where each target sample was projected, and to estimate allele frequencies as well as the effects of age, sex and PCs on the trait. The target samples were assumed to arrive sequentially. We iteratively applied the most up-to-date SNP weights, derived from all prior samples, to an incoming individual to generate a polygenic prediction, and subsequently used rtPRS-CS to update the SNP weights, after adjusting for the effects of covariates and standardizing the phenotypes and genotypes (Supplementary Figure 1). Raw PRSs for target samples estimated by rtPRS-CS were dynamically calibrated against the PRS distribution of 1000 preceding samples using top 5 genetic PCs to account for potential population substructure and mismatch between the baseline training and target samples.

The prediction accuracy of rtPRS-CS was then assessed across 10 sequential bins of the target sample, measured by contrasting the variance explained (*R*^2^) between the full model, which included both covariates (age, sex and PCs) and the PRS, and the reduced model, which included covariates only.

To assess the influence of the accuracy of the global and local shrinkage parameters on the performance of rtPRS-CS, we conducted an intermediate GWAS using the first half (five quantiles) of the UKBB target samples, and meta-analyzed the resulting GWAS with the MGBB baseline GWAS. PRS-CS-auto was then applied to the meta-GWAS to derive SNP weights and updated estimates of shrinkage parameters, which were used as a new set of starting values for running rtPRS-CS on the remaining UKBB target samples.

We compared the predictive performance of four PRSs in the UKBB holdout sample: (i) PRS derived from the baseline MGBB GWAS using PRS-CS-auto, representing the current practice for PRS construction and serving as the performance lower bound of rtPRS-CS; (ii) PRS estimated by rtPRS-CS at the end of the training process (i.e., after refining the weights using all target UKBB samples); (iii) PRS estimated by rtPRS-CS at the end of the training process, with halfway SNP weights and shrinkage parameter updates; and (iv) PRS derived from applying PRS-CS-auto to the fixed-effect meta-analysis of the MGBB baseline GWAS and GWAS of the full UKBB target sample, representing the theoretical upper bound of rtPRS-CS performance. The out-of-sample prediction accuracy was similarly measured using *R*^2^ by comparing models with and without the inclusion of the PRS.

### Schizophrenia risk prediction

We used 22 schizophrenia cohorts, collected by the Stanley Global Asia Initiatives across 7 Asian regions, totaling 27,159 schizophrenia cases and 32,336 controls (Supplementary Table 7). Sample collection and standard genetic quality control procedures using the RICOPILI pipeline^39^ have been detailed in prior publications^40^. Five cohorts, comprising a total of 4,343 cases and 7,957 controls, for which only GWAS summary statistics were available, served as the baseline training dataset. Two cohorts, JPN1 and KOR1, were imputed using 1000 Genomes Project (1KG)^29^ phase 3 data as the reference panel and were designated as holdout testing datasets. The remaining 15 cohorts were imputed using the TOPMed reference panel^41^. From these 15 cohorts, we randomly selected 1,000 cases and 1,000 controls to form the validation dataset. The remaining 21,581 cases and 23,347 controls were merged, and the order of the merged samples was randomized, which constituted the target sample. Principal component analysis (PCA) was performed using a pruned set of variants in an unrelated subset of individuals in the validation dataset. We removed variants with MAF <5%, missingness >5%, or significant deviation from Hardy-Weinberg disequilibrium (P <1e-6), and excluded individuals with genotyping rate <95%. All samples were subsequently projected onto the PC space estimated using the unrelated validation samples. These PCs were used in rtPRS-CS training and calibration.

To visualize the genetic diversity of the entire schizophrenia dataset, we combined individual-level data from all validation, target, and holdout samples. Across genotyped variants, we filtered out those with MAF <5%, missingness >5%, or significant deviation from Hardy-Weinberg disequilibrium (P <1e-10) at both the combined dataset and individual cohort levels. We also removed variants whose missingness differed by more than 0.75% between any pairwise comparison of cohorts. We additionally excluded any individuals with genotyping rate <95%. PCA was performed using a pruned set of SNPs (200kb window, *r*^2^ threshold of 0.1) in an unrelated subset of individuals, with subsequent projection of all samples onto the PC space. These PCs were only used to visualize the genetic diversity of the dataset across all cohorts and were not used in the rtPRS-CS algorithm.

We combined the GWAS summary statistics from the five baseline training cohorts through a fixed-effect meta-analysis. PRS-CS-auto was applied to the meta-GWAS to derive initial estimates of SNP weights, along with estimates of global and local shrinkage parameters. The validation sample was used to define the PC space of the dataset, where each target sample was projected as described above, and to estimate allele frequencies as well as the effects of age and PCs on schizophrenia risk. The target samples were assumed to arrive sequentially. For each incoming sample, the process of PRS calculation using weights trained on prior samples followed by updating SNP weights using rtPRS-CS was iteratively performed. Raw PRSs for target samples generated by rtPRS-CS were dynamically calibrated against the PRS distribution of 1000 preceding control samples using top 5 genetic PCs.

To evaluate the performance of rtPRS-CS, target samples with calibrated PRS were assigned back to their respective contributing cohorts. Compared to assessing the performance in the full target sample, this approach ensured that the evaluation was minimally influenced by population stratification. Within each cohort, the prediction accuracy was assessed by comparing the variance explained (*R*^2^) in the case-control status between the full model, which included both covariates (sex and top 20 in-sample PCs) and the PRS, and the reduced model, which included covariates only. For the trio and multiplex family cohorts (Supplementary Table 7) that contained closely related individuals, we pruned each cohort to a subset of unrelated individuals (kinship < 0.125) when estimating *R*^2^ to avoid potential bias. Variance explained on the observed scale was transformed to the liability scale using the method outlined in Lee et al.^42^

We compared the predictive performance of four PRSs in the two holdout cohorts: (i) PRS derived from the baseline GWAS, representing the current practice for PRS construction and the performance lower bound of rtPRS-CS; (ii) PRS estimated by rtPRS-CS at halfway through the training process; (iii) PRS estimated by rtPRS-CS at the end of the training process; and (iv) PRS derived from applying PRS-CS-auto^6^ to the fixed-effect meta-analysis of the baseline and full target sample GWASs, adjusting for sex and top 20 PCs, which represents the theoretical upper bound of rtPRS-CS performance. The out-of-sample prediction accuracy was similarly measured using variance explained by the PRS on the liability scale.

Lastly, we note that dynamically calibrated PRSs in the control group are approximately normally distributed. We therefore calculated the cutoff points corresponding to the top 20%, 10%, 5% and 2% of the standard normal distribution, and applied these cutoffs to the PRSs of target and holdout samples to identify individuals at high risk of schizophrenia.

### Ethics

The use of Mass General Brigham Biobank (MGBB) data was approved by the Mass General Brigham Institutional Review Board. Collection of the UK Biobank (UKBB) data was approved by the Research Ethics Committee of the UKBB. UKBB data used in the present work were obtained under application 32568. The use of schizophrenia cohorts of East Asian ancestry in the present work was approved by the Stanley Global Asia Initiatives. The following institutions provided ethics oversight for the collection of schizophrenia samples: Samsung Medical Center; Bio-X Institutes of Shanghai Jiao Tong University; Xi’an Jiaotong University; The Second Xiangya Hospital of Central South University; Peking University Sixth Hospital; Fujita Health University; Tokyo Metropolitan Institute of Medical Science; University Medical Center Utrecht; The University of Western Australia; The University of Indonesia; RIKEN Center for Integrative Medical Sciences; Nagoya University; Osaka University; Niigata University; Chonnam National University Hospital; and Mass General Brigham (Protocols 2014P001342 and 2011P002207). Informed consent and permission to share the data were obtained from all subjects, in compliance with the guidelines specified by the recruiting center’s institutional review board.

## Data availability

Mass General Brigham Biobank (MGBB) data are not publicly available due to privacy and ethical restrictions. De-identified data may be shared under an approved Data Use Agreement. UK Biobank (UKBB) data can be accessed under an approved application. The UKBB data used in the present study were obtained under application 32568. Data from schizophrenia cohorts are available through application to the Stanley Global Asia Initiatives. These data are subject to controlled access due to compliance requirements, participant consent and national laws. Application to access these data requires a brief research proposal that will be reviewed by the principal investigator of each cohort and, if necessary, by the respective ethics committee.

## Code availability

rtPRS-CS: https://github.com/getian107/rtPRS

PRS-CS(-auto): https://github.com/getian107/PRScs

PLINK 1.9: https://www.cog-genomics.org/plink

HAPGEN2: https://mathgen.stats.ox.ac.uk/genetics_software/hapgen/hapgen2.html

## Acknowledgements

We thank Mass General Brigham Biobank for providing genetic data and health outcomes, and all the participants and researchers of the Mass General Brigham Biobank and the UK Biobank. We thank Stanley Global Asia Initiatives for providing access to data from schizophrenia cohorts. J.D.T is supported by the Mass General Brigham Training Program in Precision and Genomic Medicine (T32HG010464). R.D. is supported by National Institute of General Medical Sciences (NIGMS) R01GM148494. H.H. acknowledges supports from National Institute of Diabetes and Digestive and Kidney Diseases (NIDDK) K01DK114379 and R01DK129364, National Institute of Mental Health (NIMH) U01MH109539 and R01MH130675, Brain and Behavior Research Foundation Young Investigator Grant (28450), the Zhengxu and Ying He Foundation, and the Stanley Center for Psychiatric Research. T.G. is supported by National Human Genome Research Institute (NHGRI) R01HG012354, U01HG011723, and NIMH R01MH130899.

## Author contributions

T.G. designed and supervised the project. T.G. developed the statistical methods with input from R.D. J.D.T. and T.G. programmed the code for rtPRS-CS. J.D.T. and T.G. conducted the simulation studies. J.D.T. performed the biobank and schizophrenia analyses. Y.C. preprocessed all schizophrenia cohorts. H.H. provided suggestions for the study design and supervised the generation and management of schizophrenia data. J.D.T. and T.G. wrote the manuscript. All the authors provided critical comments on the manuscript, and reviewed and approved the final version of the manuscript.

