## Supplementary Figures for "Real-time dynamic polygenic prediction for streaming data"

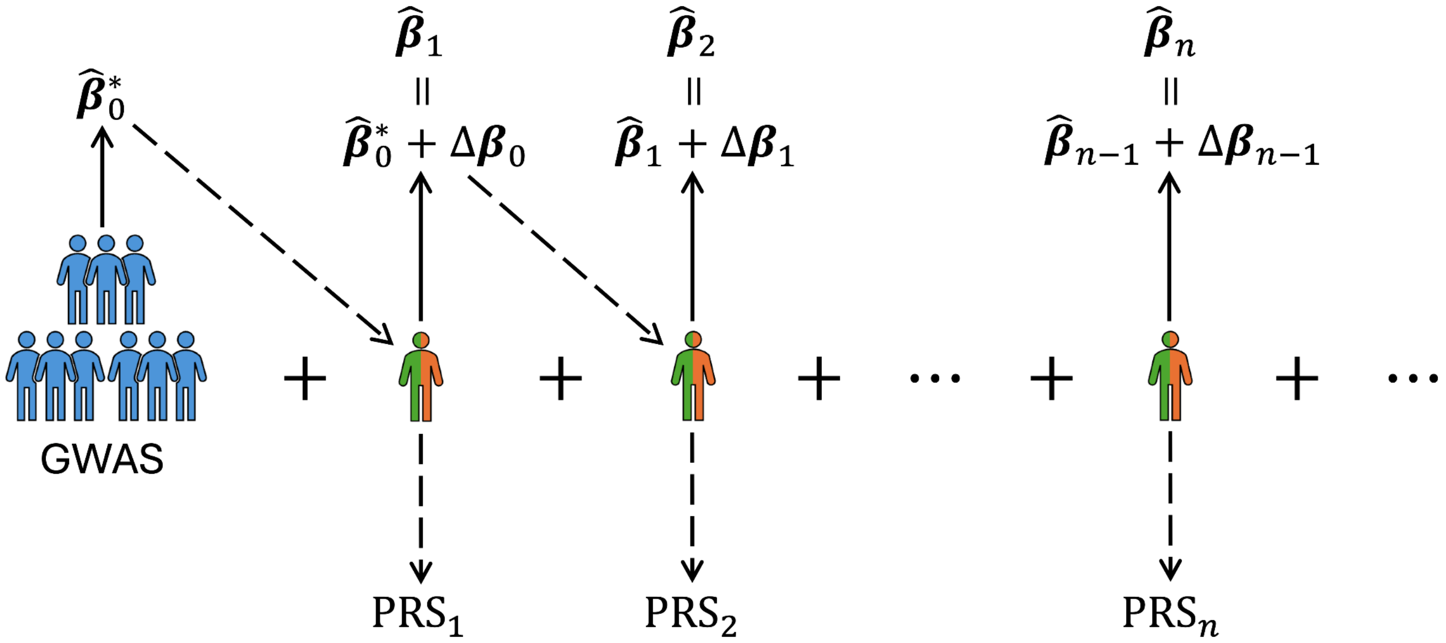


**Supplementary Figure 1: Overview of the workflow of rtPRS-CS in the present work.** Starting with the summary statistics derived from a baseline training GWAS, PRS-CS-auto is first applied to generate an initial estimate of SNP weights, denoted as $\widehat{\boldsymbol{\beta}}_0^*$, along with the global and local shrinkage parameters. Subsequently, for each newly arrived sample, the latest SNP weights are utilized to calculate the PRS, followed by the application of rtPRS-CS to update the SNP weights.

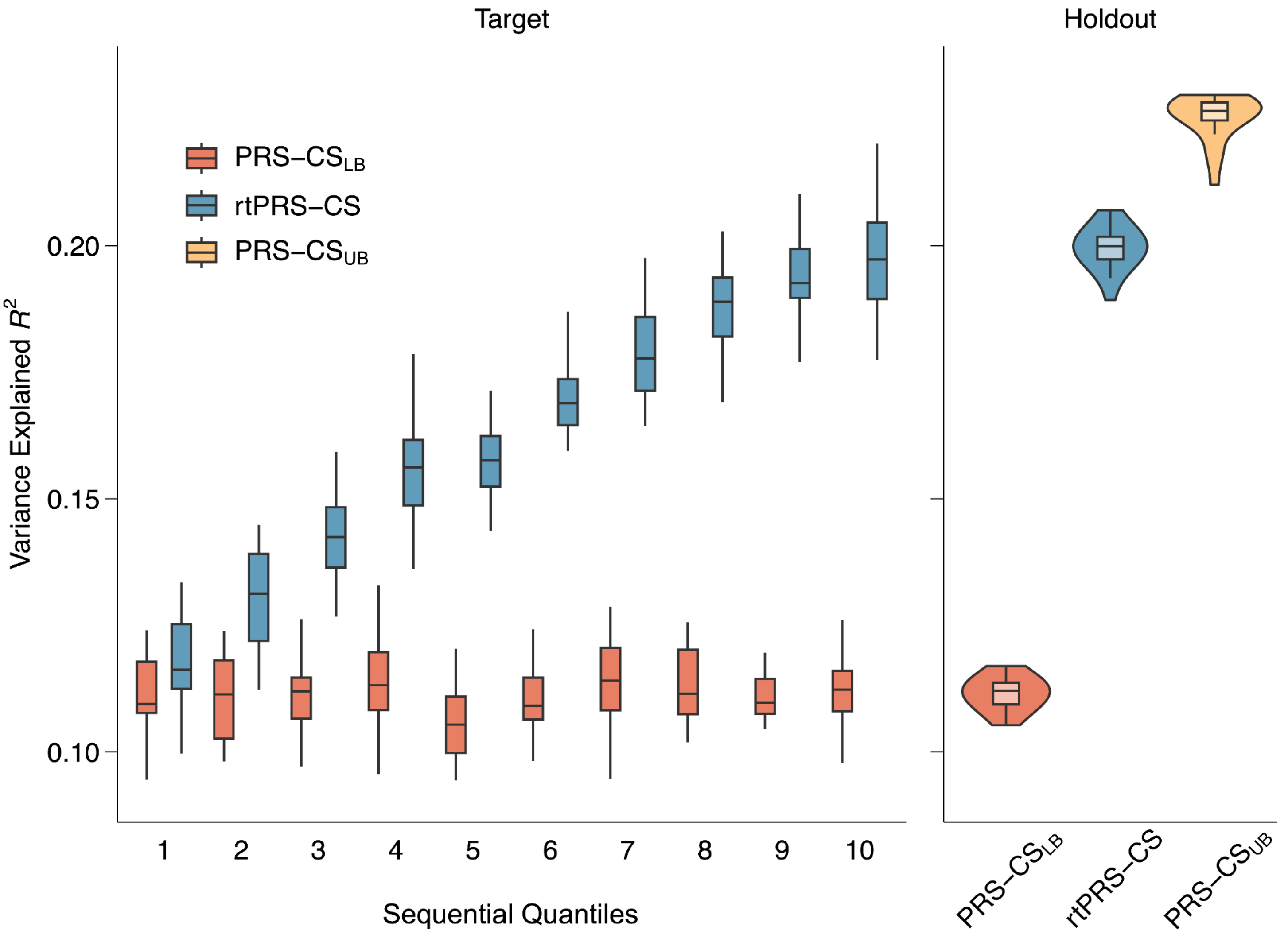


**Supplementary Figure 2: The performance of rtPRS-CS in the smaller baseline training sample size ($N$ = 25,000) simulation setting.** Left: The prediction accuracy, measured by variance explained, of the baseline PRS (red) and rtPRS-CS (blue) across 10 sequential bins of the target sample. Right: The prediction accuracy of three PRSs in the holdout sample: (i) PRS constructed from the baseline GWAS (red), representing the performance lower bound of rtPRS-CS; (ii) PRS estimated by rtPRS-CS at the end of the training process, after refining SNP weights with all target samples (blue); and (iii) PRS derived from the meta-GWAS combining baseline and target samples (yellow), representing the theoretical upper bound of the performance of rtPRS-CS. In all box plots, the middle line indicates the median across the 20 simulation replicates, and the upper and lower bound of the box indicates the 75th and 25th percentiles, respectively.

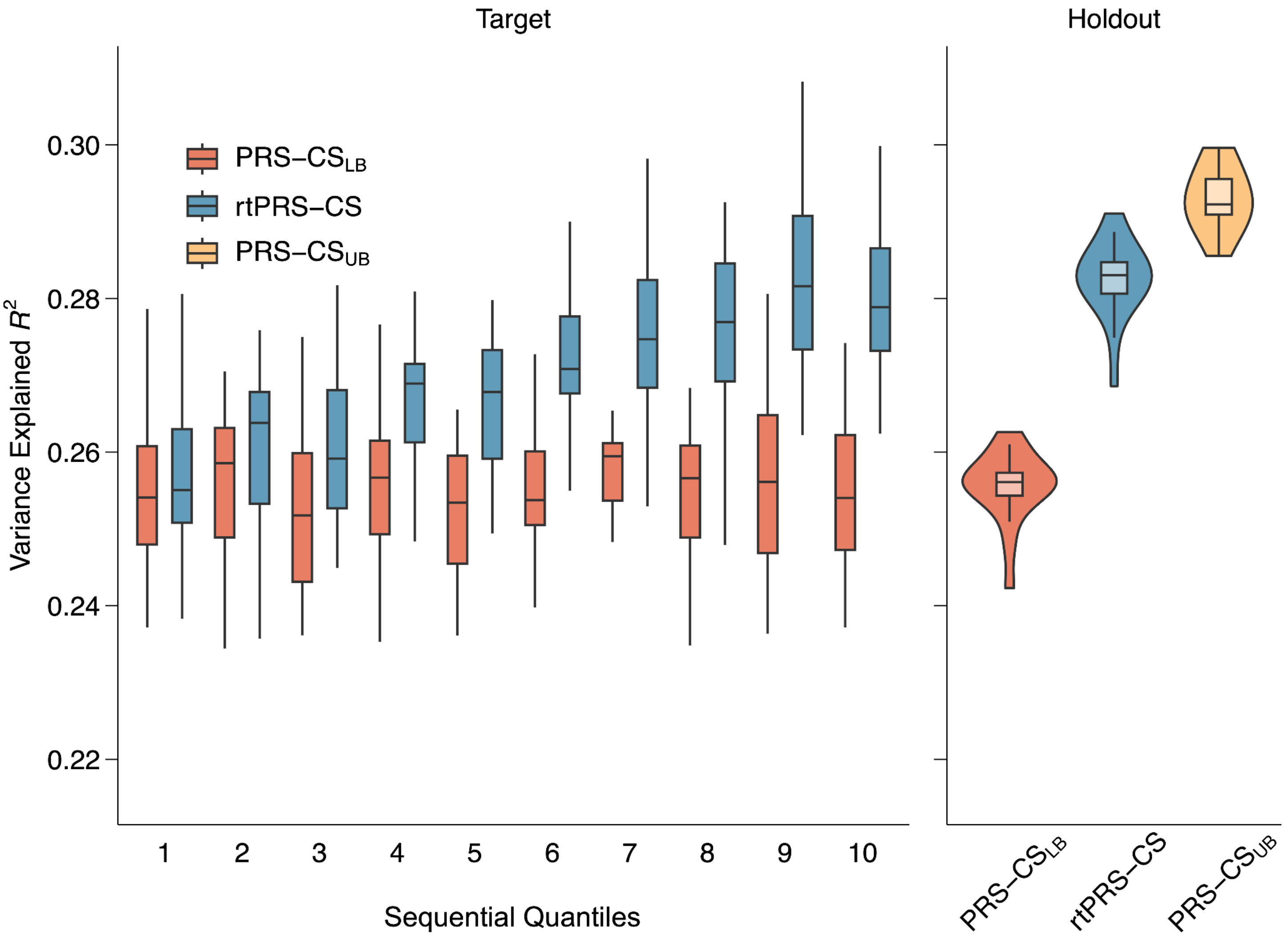


**Supplementary Figure 3: The performance of rtPRS-CS in the larger baseline training sample size (*N* = 100,000) simulation setting.** Left: The prediction accuracy, measured by variance explained, of the baseline PRS (red) and rtPRS-CS (blue) across 10 sequential bins of the target sample. Right: The prediction accuracy of three PRSs in the holdout sample: (i) PRS constructed from the baseline GWAS (red), representing the performance lower bound of rtPRS-CS; (ii) PRS estimated by rtPRS-CS at the end of the training process, after refining SNP weights with all target samples (blue); and (iii) PRS derived from the meta-GWAS combining baseline and target samples (yellow), representing the theoretical upper bound of the performance of rtPRS-CS. In all box plots, the middle line indicates the median across the 20 simulation replicates, and the upper and lower bound of the box indicates the 75th and 25th percentiles, respectively.

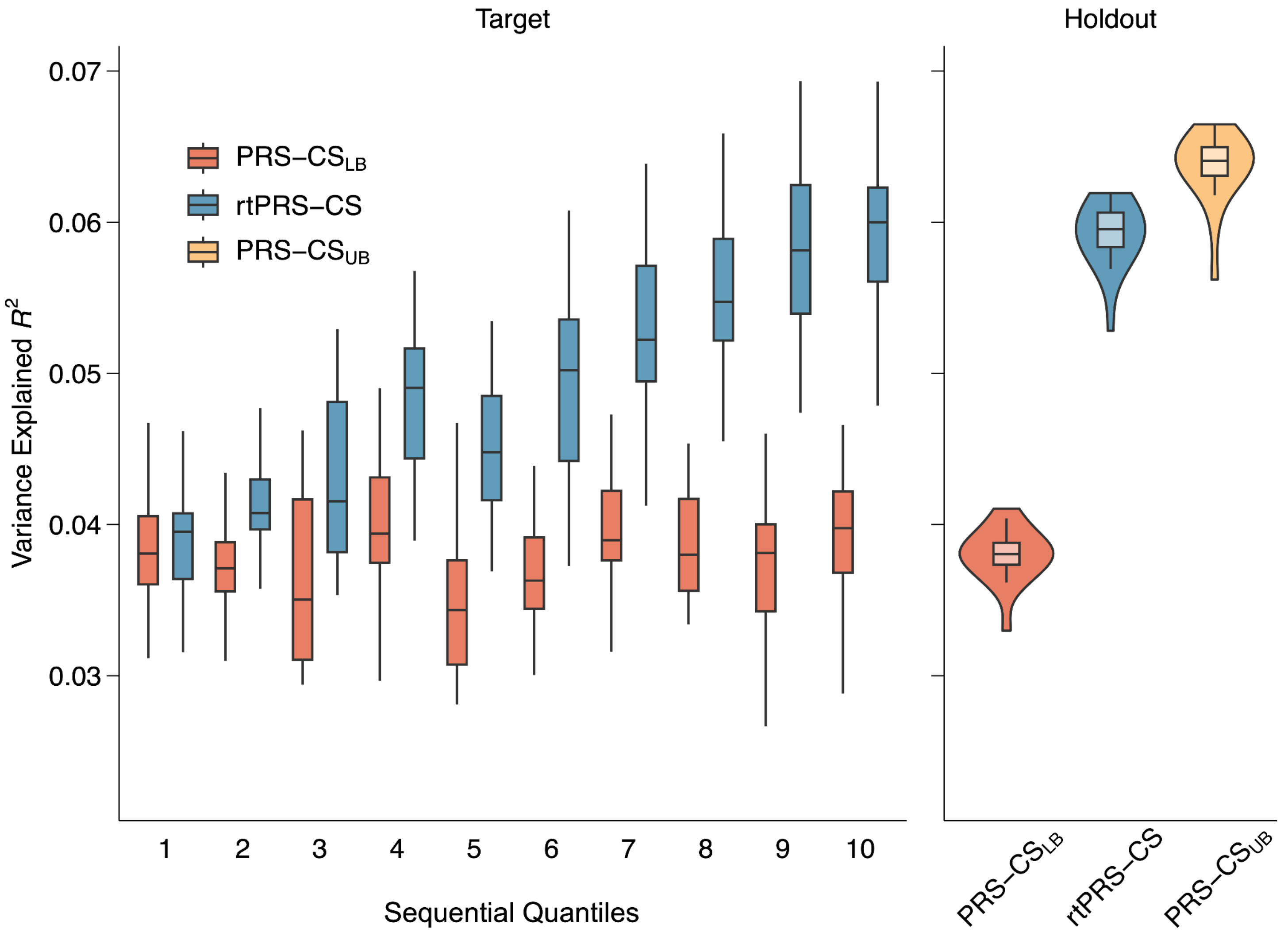


**Supplementary Figure 4: The performance of rtPRS-CS in the lower heritability ($h^2$ = 0.2) simulation setting.** Left: The prediction accuracy, measured by variance explained, of the baseline PRS (red) and rtPRS-CS (blue) across 10 sequential bins of the target sample. Right: The prediction accuracy of three PRSs in the holdout sample: (i) PRS constructed from the baseline GWAS (red), representing the performance lower bound of rtPRS-CS; (ii) PRS estimated by rtPRS-CS at the end of the training process, after refining SNP weights with all target samples (blue); and (iii) PRS derived from the meta-GWAS combining baseline and target samples (yellow), representing the theoretical upper bound of the performance of rtPRS-CS. In all box plots, the middle line indicates the median across the 20 simulation replicates, and the upper and lower bound of the box indicates the 75th and 25th percentiles, respectively.

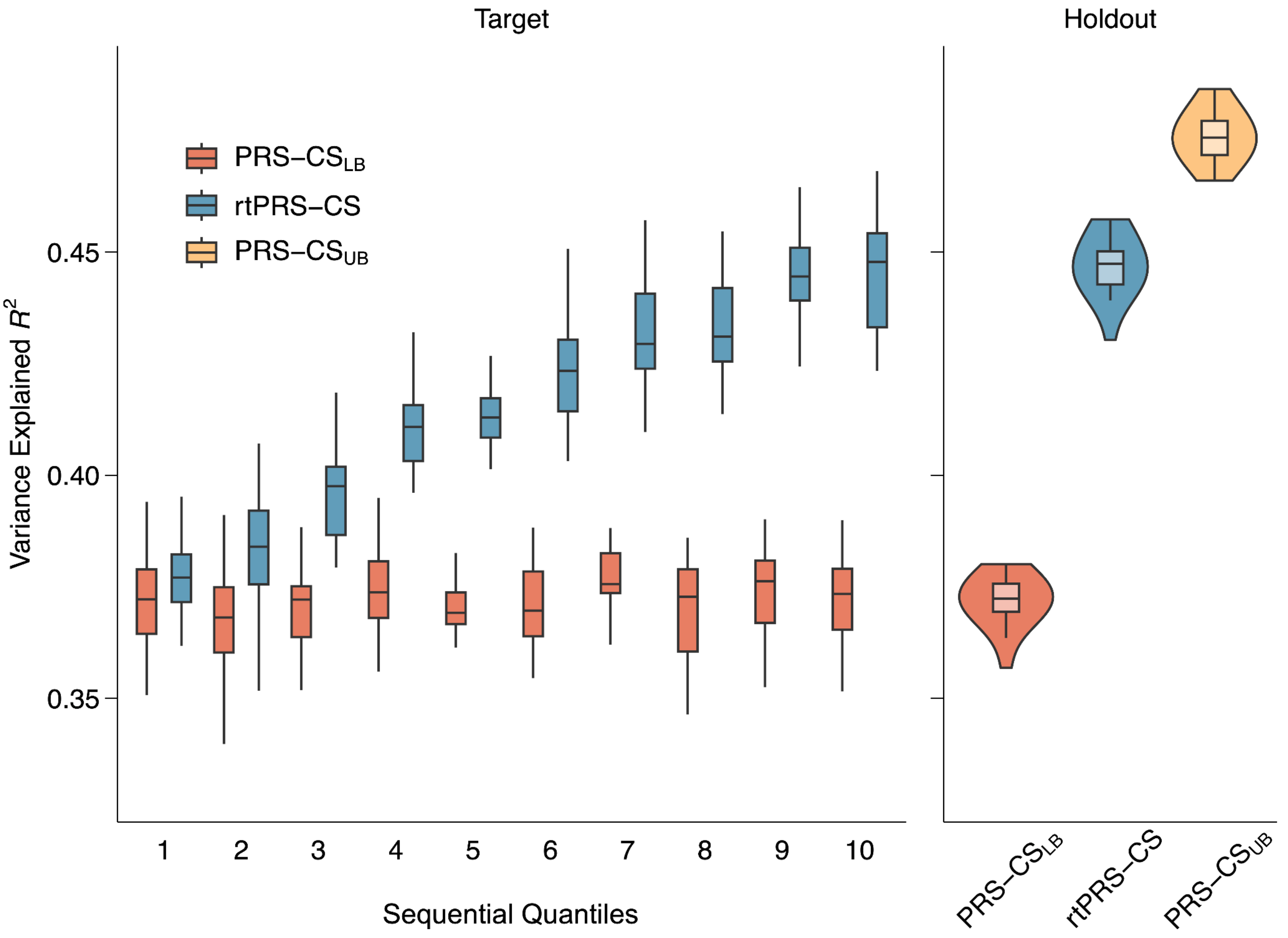


**Supplementary Figure 5: The performance of rtPRS-CS in the higher heritability ($h^2$ = 0.8) simulation setting.** Left: The prediction accuracy, measured by variance explained, of the baseline PRS (red) and rtPRS-CS (blue) across 10 sequential bins of the target sample. Right: The prediction accuracy of three PRSs in the holdout sample: (i) PRS constructed from the baseline GWAS (red), representing the performance lower bound of rtPRS-CS; (ii) PRS estimated by rtPRS-CS at the end of the training process, after refining SNP weights with all target samples (blue); and (iii) PRS derived from the meta-GWAS combining baseline and target samples (yellow), representing the theoretical upper bound of the performance of rtPRS-CS. In all box plots, the middle line indicates the median across the 20 simulation replicates, and the upper and lower bound of the box indicates the 75th and 25th percentiles, respectively.

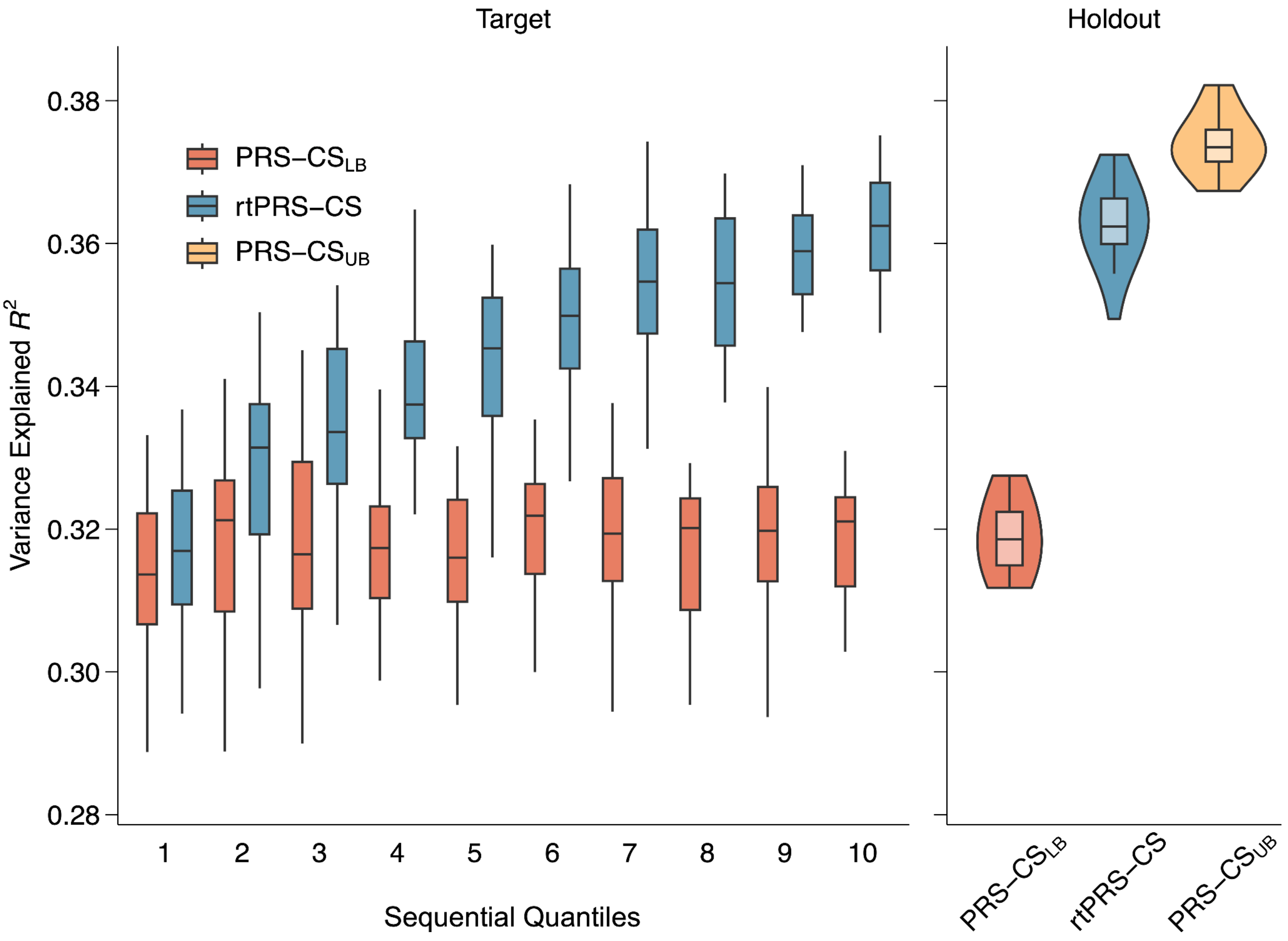


**Supplementary Figure 6: The performance of rtPRS-CS in the lower polygenicity ($\pi$ = 0.1%) simulation setting.** Left: The prediction accuracy, measured by variance explained, of the baseline PRS (red) and rtPRS-CS (blue) across 10 sequential bins of the target sample. Right: The prediction accuracy of three PRSs in the holdout sample: (i) PRS constructed from the baseline GWAS (red), representing the performance lower bound of rtPRS-CS; (ii) PRS estimated by rtPRS-CS at the end of the training process, after refining SNP weights with all target samples (blue); and (iii) PRS derived from the meta-GWAS combining baseline and target samples (yellow), representing the theoretical upper bound of the performance of rtPRS-CS. In all box plots, the middle line indicates the median across the 20 simulation replicates, and the upper and lower bound of the box indicates the 75th and 25th percentiles, respectively.

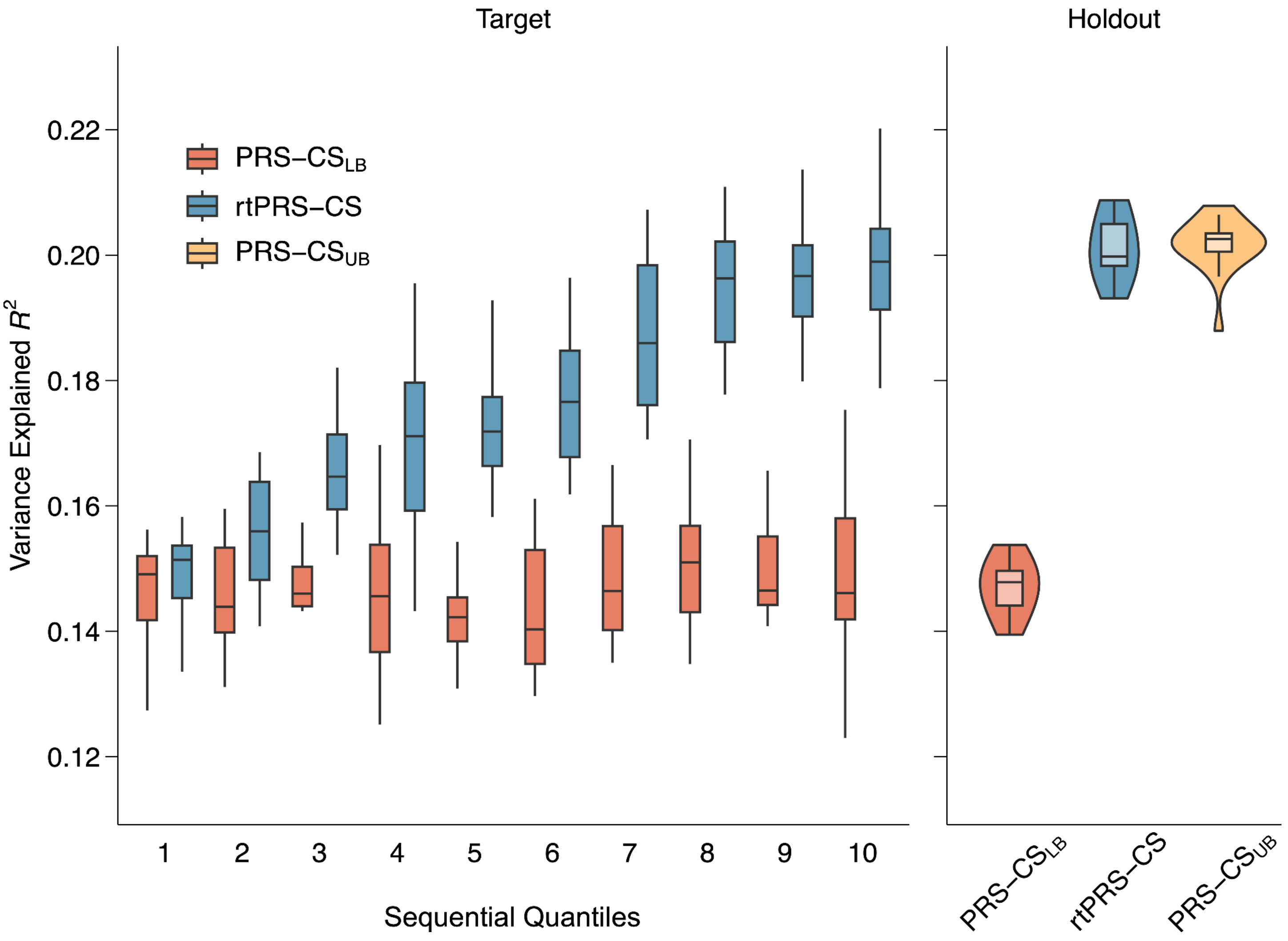


**Supplementary Figure 7: The performance of rtPRS-CS in the higher polygenicity ($\pi$ = 10%) simulation setting.** Left: The prediction accuracy, measured by variance explained, of the baseline PRS (red) and rtPRS-CS (blue) across 10 sequential bins of the target sample. Right: The prediction accuracy of three PRSs in the holdout sample: (i) PRS constructed from the baseline GWAS (red), representing the performance lower bound of rtPRS-CS; (ii) PRS estimated by rtPRS-CS at the end of the training process, after refining SNP weights with all target samples (blue); and (iii) PRS derived from the meta-GWAS combining baseline and target samples (yellow), representing the theoretical upper bound of the performance of rtPRS-CS. In all box plots, the middle line indicates the median across the 20 simulation replicates, and the upper and lower bound of the box indicates the 75th and 25th percentiles, respectively.

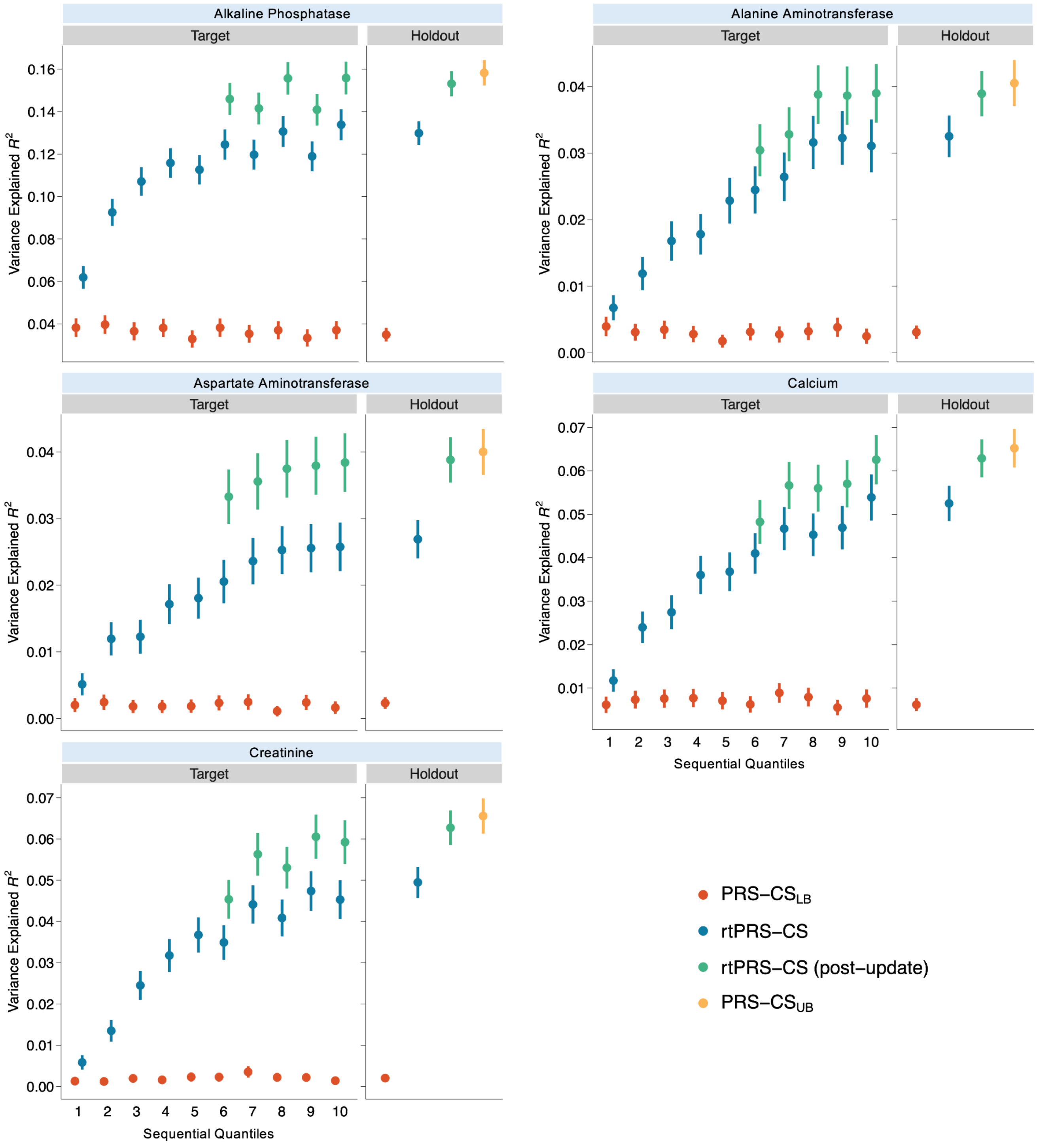


**Supplementary Figure 8: The performance of rtPRS-CS for representative quantitative traits in biobanks.** For each trait, Left: The prediction accuracy, measured by variance explained, of the baseline PRS (red), rtPRS-CS (blue), and rtPRS-CS with halfway training GWAS update (green), across 10 sequential bins of the UK Biobank target sample. Right: The prediction accuracy of four PRSs in the UK Biobank holdout sample: (i) PRS constructed from the baseline GWAS (red), representing the performance lower bound of rtPRS-CS; (ii) PRS estimated by rtPRS-CS at the end of the training process, after refining SNP weights with all target samples (blue); (iii) PRS estimated by rtPRS-CS with halfway update of the GWAS (green); and (iv) PRS derived from the meta-GWAS combining baseline and target samples (yellow), representing the theoretical upper bound of the performance of rtPRS-CS. In all plots, the error bars represent 95% confidence intervals.

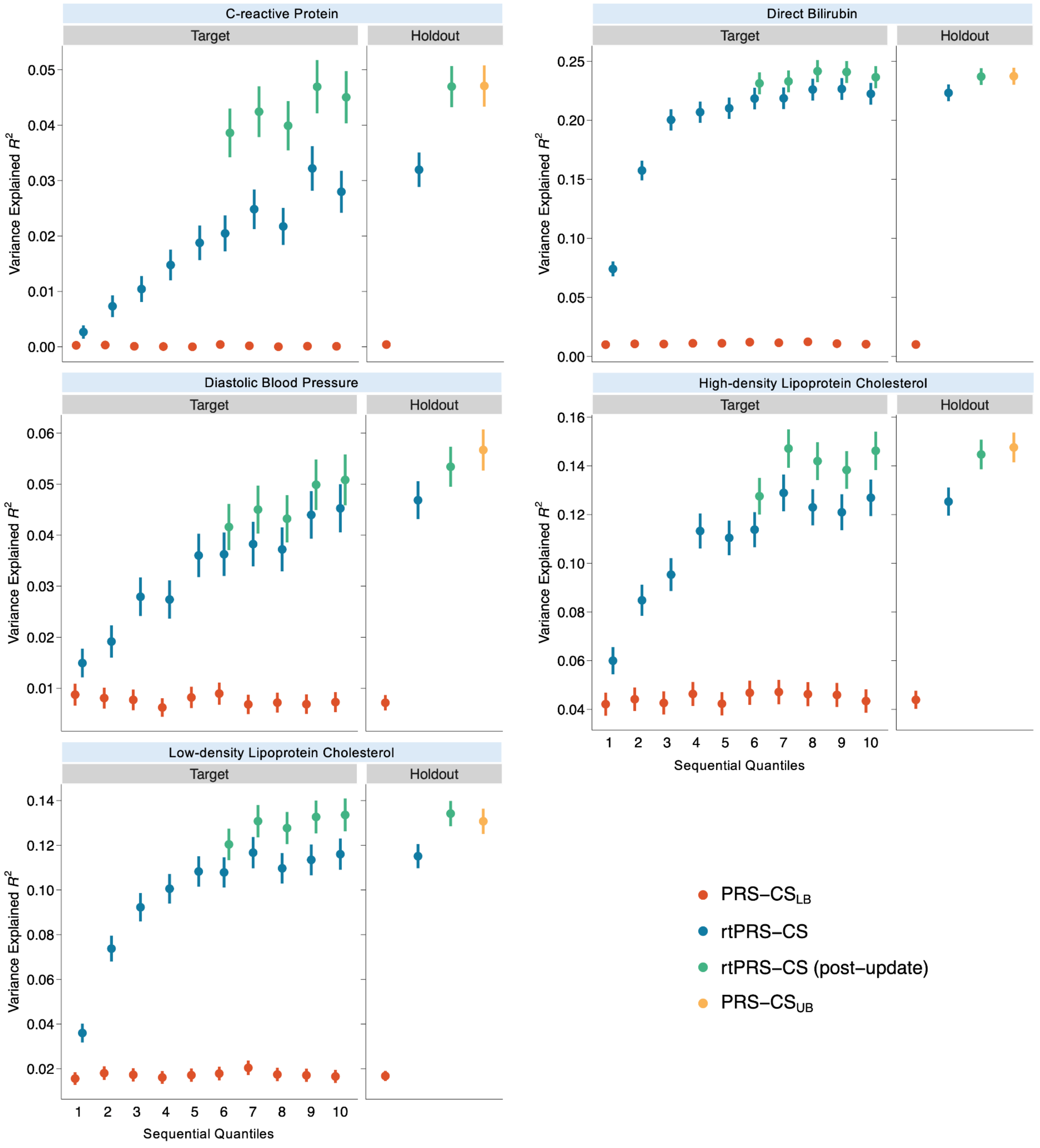


**Supplementary Figure 9: The performance of rtPRS-CS for representative quantitative traits in biobanks.** For each trait, Left: The prediction accuracy, measured by variance explained, of the baseline PRS (red), rtPRS-CS (blue), and rtPRS-CS with halfway training GWAS update (green), across 10 sequential bins of the UK Biobank target sample. Right: The prediction accuracy of four PRSs in the UK Biobank holdout sample: (i) PRS constructed from the baseline GWAS (red), representing the performance lower bound of rtPRS-CS; (ii) PRS estimated by rtPRS-CS at the end of the training process, after refining SNP weights with all target samples (blue); (iii) PRS estimated by rtPRS-CS with halfway update of the GWAS (green); and (iv) PRS derived from the meta-GWAS combining baseline and target samples (yellow), representing the theoretical upper bound of the performance of rtPRS-CS. In all plots, the error bars represent 95% confidence intervals.

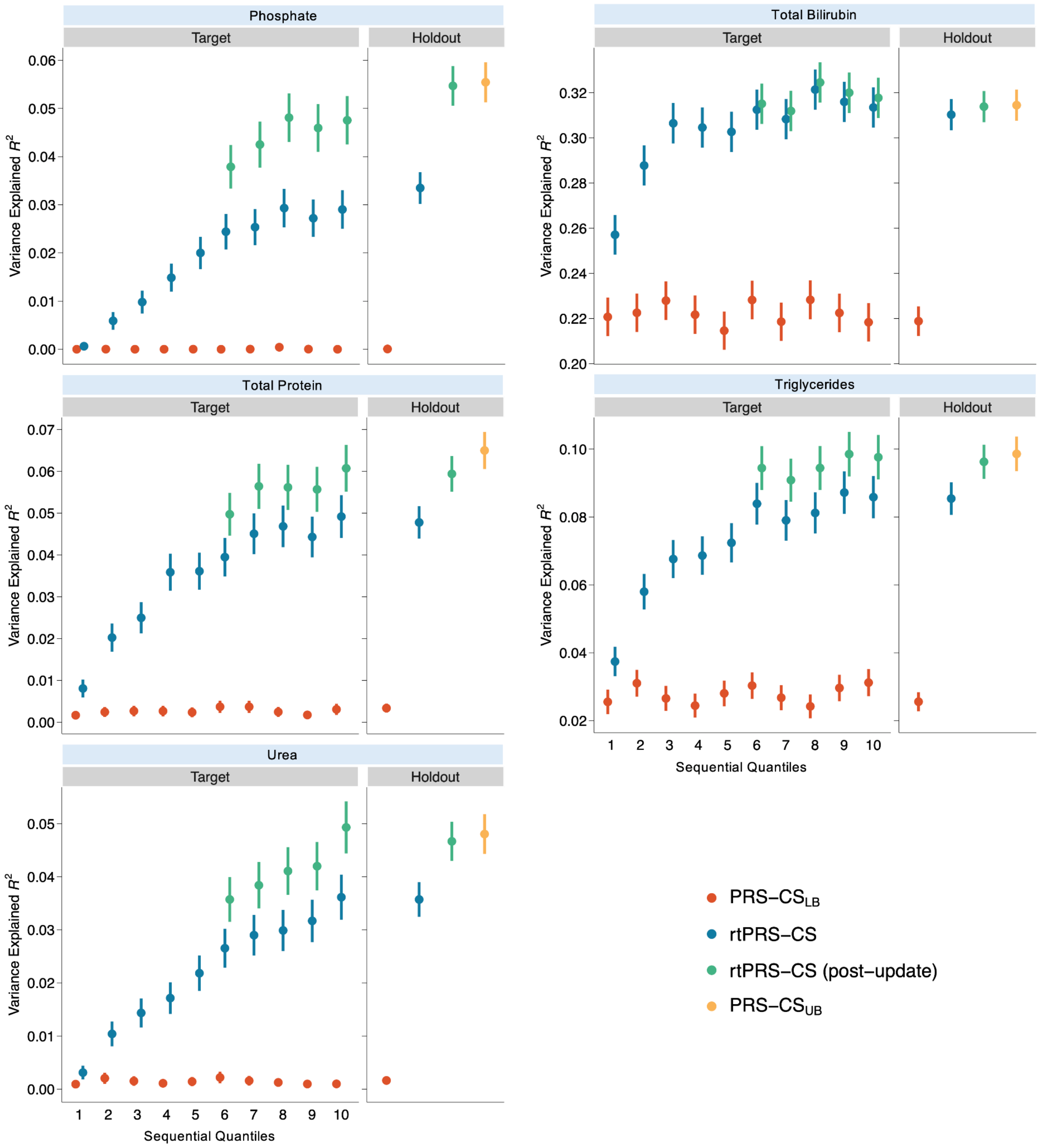


**Supplementary Figure 10: The performance of rtPRS-CS for representative quantitative traits in biobanks.** For each trait, Left: The prediction accuracy, measured by variance explained, of the baseline PRS (red), rtPRS-CS (blue), and rtPRS-CS with halfway training GWAS update (green), across 10 sequential bins of the UK Biobank target sample. Right: The prediction accuracy of four PRSs in the UK Biobank holdout sample: (i) PRS constructed from the baseline GWAS (red), representing the performance lower bound of rtPRS-CS; (ii) PRS estimated by rtPRS-CS at the end of the training process, after refining SNP weights with all target samples (blue); (iii) PRS estimated by rtPRS-CS with halfway update of the GWAS (green); and (iv) PRS derived from the meta-GWAS combining baseline and target samples (yellow), representing the theoretical upper bound of the performance of rtPRS-CS. In all plots, the error bars represent 95% confidence intervals.
